# COVID-19 Associated Mucormycosis: Scoping Review Protocol

**DOI:** 10.1101/2021.05.30.21258068

**Authors:** Salman Hussain, Harveen Baxi, Abanoub Riad, Jitka Klugarová, Radim Líčeník, Miloslav Klugar

**Affiliations:** Czech National Centre for Evidence-Based Healthcare and Knowledge Translation (Cochrane Czech Republic, Czech EBHC: JBI Centre of Excellence, Masaryk University GRADE Centre), Institute of Biostatistics and Analyses, Faculty of Medicine, Masaryk University, Brno, Czech Republic; Independent Researcher, New Delhi, India; Department of Public Health, Faculty of Medicine, Masaryk University, 62500 Brno, Czech Republic

**Keywords:** COVID-19, Fungal Infection, Mucormycosis, Scoping Review, Systematic Review

## Abstract

**Background and Objective:** Mucormycosis, a serious and rare fungal infection, has occurred concurrently in COVID-19 patients globally. Mucormycosis is associated with a high risk of all-cause mortality, with mortality depending on body site infected, fungus type, and the patient’s overall condition. This deadly fungal infection is quite difficult and expensive to treat. Therefore, this scoping review aims to map all the empirical evidence on the COVID-19 associated mucormycosis with a special focus on clinical presentation, treatment, and patient outcomes.

**Methods:** The proposed scoping review will be developed by adhering to the JBI methodology for scoping reviews and will be reported as per the Preferred Reporting Items for Systematic Reviews and Meta-analyses for Scoping Reviews (PRISMA-ScR). A detailed search strategy has already been utilized to locate both published and unpublished studies. Literature search was carried out in three phases. PubMed, Scopus, Cochrane, Google scholar was searched. We will include all those studies which are presenting mucormycosis cases in COVID-19 positive patients. Data will be extracted in a pre-designed data extraction sheet and risk of bias will be assessed using JBI critical appraisal tool. The primary outcome will be to summarize the clinical presentation, treatment, and patient outcomes in COVID-19 associated mucormycosis. Data will be presented in tabular form.

**OSF registration number:** osf.io/438sm

## Introduction

The COVID-19 outbreak, caused by severe-acute-respiratory-syndrome-coronavirus-2 (SARS-CoV-2), has infected more than 164 million people globally with approximately 3.4 million deaths.[1] COVID-19 patients often have several co-morbidities including diabetes.[2] Ample evidence has proved patients with co-morbidities to be at a higher risk of ICU admissions and mortality than those without comorbidities.[3-5] COVID-19 patients in severe stages (admitted to ICUs), are prescribed high-doses of steroids as a life saving measure.[6] Steroids, supress the immune system (decrease in CD4 + T and CD8 + T cells) to fight against the inflammation caused by the virus, thereby creating a favourable environment for other opportunistic infections.[6, 7] Mucormycosis, a serious and rare fungal infection, has occurred concurrently in COVID-19 cases globally.[8] Mucormycosis, also known as black fungus, is an invasive fungal infection most commonly caused by fungi–Rhizpous.[9] This deadly fungal infection is quite difficult and expensive to treat. At the moment more detailed evidence on the clinical presentation, treatment and patient outcomes is required. The preliminary search for existing scoping reviews was performed in Epistemonikos, PROSPERO, Open Science Framework, Cochrane Library and JBI Evidence Synthesis and no previous scoping reviews were identified.

### Objective

This scoping review will be conducted to map all empirical evidence on the COVID-19 associated mucormycosis, with a special focus on clinical presentation, treatment, and patient outcomes.

## Methods

The proposed scoping review will be developed by adhering to the JBI methodology for scoping reviews and will be reported as per the Preferred Reporting Items for Systematic Reviews and Meta-analyses for Scoping Reviews (PRISMA-ScR).[10, 11]

### Search strategy

A detailed search strategy has already been utilized to locate both published and unpublished studies. Literature search was carried out in three phases. First, an initial limited search was undertaken in PubMed, using keywords and index terms related to COVID-19, SARA-CoV2, mucormycosis, and black fungus. An analysis of the text words contained in the title and abstract and the index terms used to describe the articles was then followed. A second search using all identified keywords and index terms was conducted in all included databases (PubMed, Scopus, Cochrane, Google scholar). Refer Appendix 1 for the detailed search strategy. Third, the reference lists of all studies that met the inclusion criteria were checked for other additional records followed by citation tracking. Gray literatures were located using Cochrane COVID-19 study register, bioRxiv, and medRxiv. COVIDENCE will be use to complete this scoping review (Covidence systematic review software, Veritas Health Innovation, Melbourne, Australia. Available at www.covidence.org).

### Study selection

Two independent reviewers (SH and HB) will be screening all the retrieved articles in alignment to the inclusion criteria. In the initial screening phase, articles will be selected based on the title and abstract scanning. Then, in the second phase, full-text screening will be performed for the final inclusion of articles in the scoping review. PRISMA flow diagram will depict each stage of the study selection process.

### Inclusion criteria

#### Participants

Patients with confirmed COVID-19 (RT-PCR) and mucormycosis (either histologically or microbiologically confirmed)

#### Concept

This review will include all studies that describe the clinical presentation, treatment, and patient outcomes.

#### Context

We will focus on the clinical studies discussing the clinical presentation, treatment, and patient outcomes in COVID-19 patients with mucormycosis.

#### Type of sources

We will include descriptive studies (case-report, case-series, cross-sectional) and epidemiological studies.

We will exclude:

- Non-English language studies
- Studies with no clear information about mucormycosis and other criteria’s
- Systematic reviews, narrative reviews, editorials, opinions, correspondence, and study protocols will be excluded

### Assessment of risk of bias

The methodological quality of the selected studies will be evaluated using the Jonna Briggs Institute (JBI) critical appraisal tool. An appropriate JBI risk-of-bias tool will be adapted according to the study design of the studies qualified for inclusion in this scoping review. Depending on the response of each domain of the tool, the study will be classified as high, low or unclear risk of bias.

### Data analysis and presentation

Two reviewers (SH and HB) will independently extract the data in a pre-designed data extraction template. Following information will be extracted from all eligible studies qualified for inclusion: Study author, year of publication, country, study design, characteristics of the population (age, sex), sample size, co-morbidities, treatment for COVID-19, symptoms of mucormycosis, diagnosis of mucormycosis, identification of fungal species, treatment for mucormycosis, and patient outcomes. Data will be descriptively presented for each outcome.

## Supporting information

Search Strategy

## Data Availability

The data that support the findings of this study are available from the corresponding author upon reasonable request.

