## Supplementary material for "COVID-19 Associated Mucormycosis: Scoping Review Protocol": Search Strategy

**Appendix I**

1. coronavirus*
2. coronovirus*
3. coronavirinae*
4. Coronavirus*
5. Coronovirus*
6. Wuhan*
7. Hubei*
8. 2019-nCoV
9. 2019nCoV
10. nCoV2019
11. nCoV-2019
12. COVID-19
13. COVID19
14. HCoV-19
15. 2019 novel*
16. SARS-CoV-2
17. SARSCoV-2
18. SARSCoV2
19. SARS-CoV2
20. SARSCov19
21. SARS-Cov19
22. SARS-Cov-19
23. OR/1-22
24. mucormycosis
25. black fungus
26. mucor*
27. mucorales
28. zygomycosis
29. zygomy*
30. rhizopus
31. rhizomucor
32. absidia
33. apophysomyces
34. cokeromyces
35. OR/24-34
36. AND/23,35
